# Linking the plasma proteome to genetics in individuals from continental Africa provides insights into type 2 diabetes pathogenesis

**DOI:** 10.1101/2024.09.16.24313728

**Authors:** Opeyemi Soremekun, Young-Chan Park, Mauro Tutino, Allan Kalungi, N. William Rayner, Moffat Nyirenda, Segun Fatumo, Eleftheria Zeggini

## Abstract

Individuals of African ancestry remain largely underrepresented in genetic and proteomic studies. Here, we measure the levels of 2,873 proteins using the Olink proximity extension assay in plasma samples from 163 individuals with type 2 diabetes (T2D) or prediabetes and 362 normoglycemic controls from the Ugandan population for the first time. We identify 88 differentially expressed proteins between the two groups and 208 proteins associated with cardiometabolic traits. We link genome-wide data to protein expression levels and construct the first protein quantitative trait locus (pQTL) map in this population. We identify 399 independent associations with 346 (86.7%) cis*-*pQTLs and 53 (13.3%) trans*-*pQTLs. 16.7% of the cis-pQTLs and all of the trans-pQTLs have not been previously reported in African-ancestry individuals. Of these, 37 pQTLs have not been previously reported in any population. We find evidence for colocalization between a pQTL for SIRPA and T2D genetic risk. Mendelian randomization analysis identified 20 proteins causally associated with T2D. Our findings reveal proteins causally implicated in the pathogenesis of T2D, which may be leveraged for personalized medicine tailored to African-ancestry individuals.

## Main

Type 2 diabetes (T2D) is becoming a major public health concern in Africa, congruent with the complex interplay of genetic, environmental and socio-economic factors ^1–3^. The shift from traditional to urbanized and sedentary lifestyles, accompanied by dietary changes, has contributed to this increasing prevalence^4–6^. According to the International Diabetes Federation (IDF), it is predicted that, globally, people with T2D will rise by 51% reaching 700.2 million by 2045 from 463 million in 2019 ^7^. A significant increase of 143% is anticipated in Africa, with numbers expected to rise from 19.4 million in 2019 to 47.1 million in 2045 ^7^. Hemoglobin A1c (HbA1c), also known as glycated hemoglobin ^8^, provides an estimate of the blood sugar level over a period of two to three months by measuring the percentage of hemoglobin with attached glucose ^9,10^. An HbA1c level of 6.5% or higher on two separate tests typically indicates diabetes. Levels between 5.7% and 6.4% suggest prediabetes, and values below 5.7% are considered normal ^11^.

Proteins drive many biological functions, can be used as biomarkers of disease onset and progression, and are the primary targets of drug therapies. With advancement in technologies, high-throughput quantification of circulating proteins on an epidemiological scale is now possible. Combining proteomic and genomic data for blood-based pQTLs has led to the identification of hundreds of associations between genetic variants and protein levels ^12–16^. A fraction of African-ancestry individuals in diaspora have been studied in proteomics studies to date ^15,17^, with continental Africans largely underrepresented.

To address this, we have measured 2873 proteins using the Olink PEA Explore assay in plasma samples of 163 individuals with prediabetes or type 2 diabetes (cases) (defined as HbA1c > 5.7%) as well as 362 normoglycemic controls (defined as HbA1c < 5.7%) (Table 1). We have performed differential protein expression analysis between the two groups, and have undertaken proteomic genetic association analysis to identify sequence variants influencing protein levels. We have subsequently examined the role of the identified pQTLs in type 2 diabetes using colocalization and Mendelian randomization analysis.

**Table 1:** Clinical Charateristics of study participants.

|  | Cases | Controls |
| --- | --- | --- |
| Number of participants (n, %) | 163(31.05%) | 362(68.95%) |
| Age (year) (mean ± SD) | 49.82 ± 18.5 | 50.39 ± 17.76 |
| Male (n, %) | 47(28.83%) | 139(38.40%) |
| Female (n, %) | 116(71.17%) | 223(61.60%) |
| BMI (kg/m<sup>2</sup>) | 23.4 ± 4.68 | 22.06 ± 4.35 |
| HbA1c (%) | 6.46 ± 1.24 | 5.13 ± 0.48 |

First, we studied the association between protein levels and cardiometabolic traits measured in the Ugandan cohort (supplementary 1). A total of 208 proteins were associated with HbA1c, 42 with HDL, and 46 with LDL at a false discovery rate (FDR) of 5% (Figure 1). Some of the associations, such as ERCC1 found to be associated with HbA1c (P_adj_ = 6.77×10^−7^) and HDL (P_adj_ = 1.91×10^−2^), have been shown to affect glucose intolerance in a progeroid-deficient animal model causing an autoinflammatory response that leads to fat loss and insulin resistance ^18^. PTPN9 was also found to be associated with HbA1c (2.96×10^−6^); this protein belongs to a family of protein tyrosine phosphatases (PTN) that are known to be involved in cellular insulin resistance associated with T2D ^19,20^. Due to the role PTPN9 plays in insulin resistance, it is currently being investigated as a potential diabetes drug target ^21,22^.

**Figure 1:**
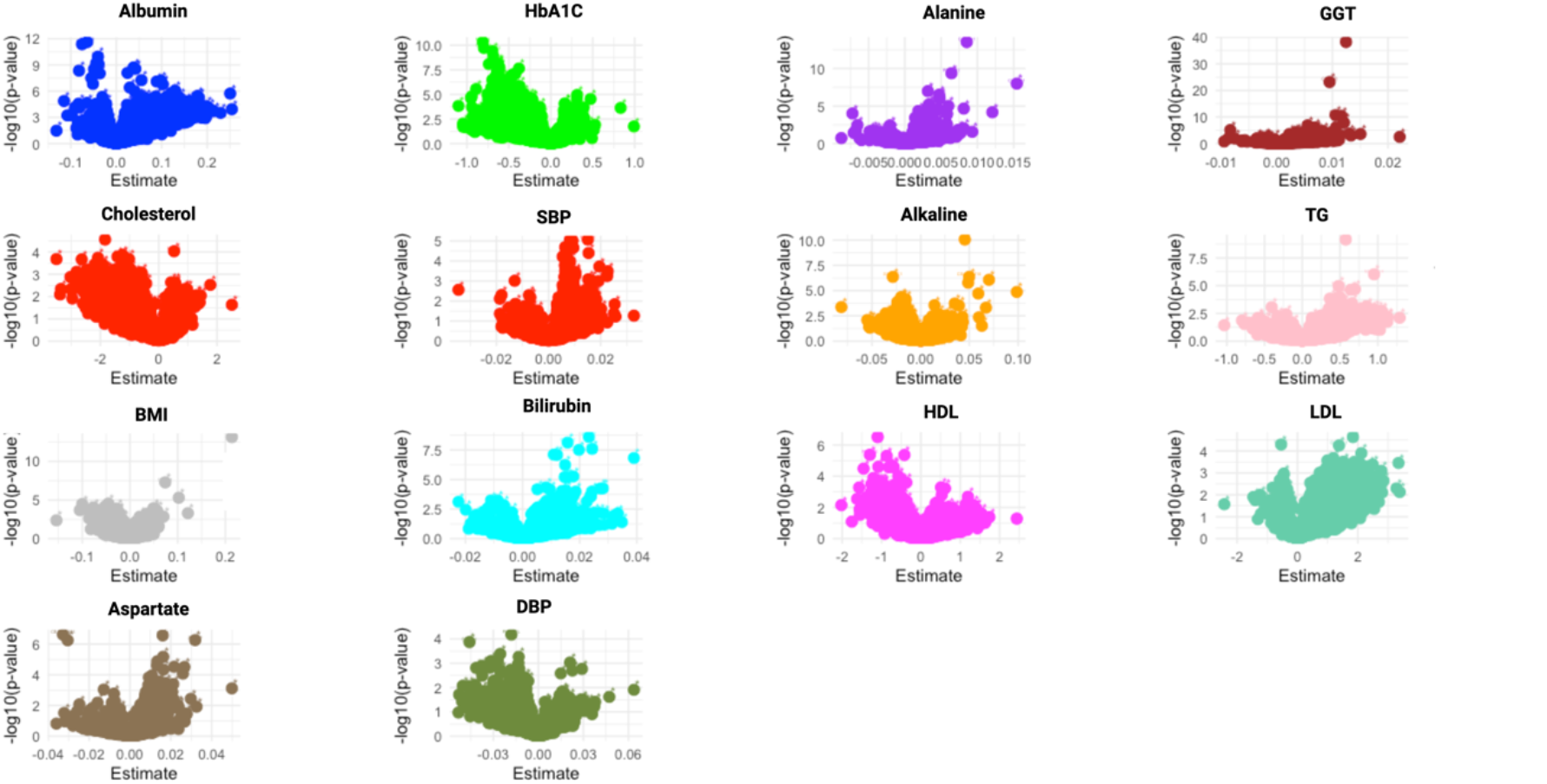
Association of protein levels with clinical traits. The y axis represents the FDR-adjusted -log 10(p-value) of the association and the x axis of each plot represents the effect size estimated with linear regression. [Body mass index (BMI), Systolic blood pressure (SBP), Diastolic blood pressure (DBP), High density lipoproteins (HDL), Gamma glutamate transferase (GGT), Triglycerides (TG), Low density lipoproteins (LDL)].

Next, we sought to identify differentially expressed protein (DEP) levels between cases and controls. Using a set of 2873 unique proteins, DEPs were defined based on a 2-fold change (|log2FC| > 0.5) in expression levels at a false discovery rate (FDR) of 5%. This led to the identification of 88 DEPs. Among these, 57 were found to be significantly upregulated, with log2 fold changes ranging from 0.50 to 1.18, while 31 proteins were downregulated with log2 fold changes between -0.51 to -1.17 (Figure 2A), (supplementary 2). EDIL3 ((EGF-like repeats and discoidin I-like domains 3), associated with processes such as cell adhesion, migration and vascular development, showed the most significant upregulation with an adjusted p-value of 1.2×10^−13^. EDIL3 has been shown to be differentially expressed in adipose tissue of insulin-resistant and insulin sensitive individuals ^23,24^, and to be involved in angiogenesis ^25–27^. Impaired angiogenesis has been implicated in the progression of diabetic retinopathy and nephropathy ^28,29^. Studies also indicate that EDIL3 is involved in inflammatory responses, which are known to contribute to insulin resistance, a hallmark of T2D^30^. LPCAT2 (Lysophosphatidylcholine Acyltransferase 2) was identified as the most significantly downregulated protein with Padj = 4.41×10^−14^. LPCAT2 plays a critical role in lipid metabolism, specifically in the remodeling of phospholipids in cell membranes ^31,32^. Downregulation of LPCAT2 may lead to changes in membrane lipids that contribute to impaired insulin signaling. The DEPs were primarily enriched in GO terms such as chemokine receptor binding, and chemokine and cytokine activity (Figure 2B, supplementary 3). For the first time, EDIL3 and LPCAT2 are identified to be differentially expressed between diabetes cases and controls.

**Figure 2:**
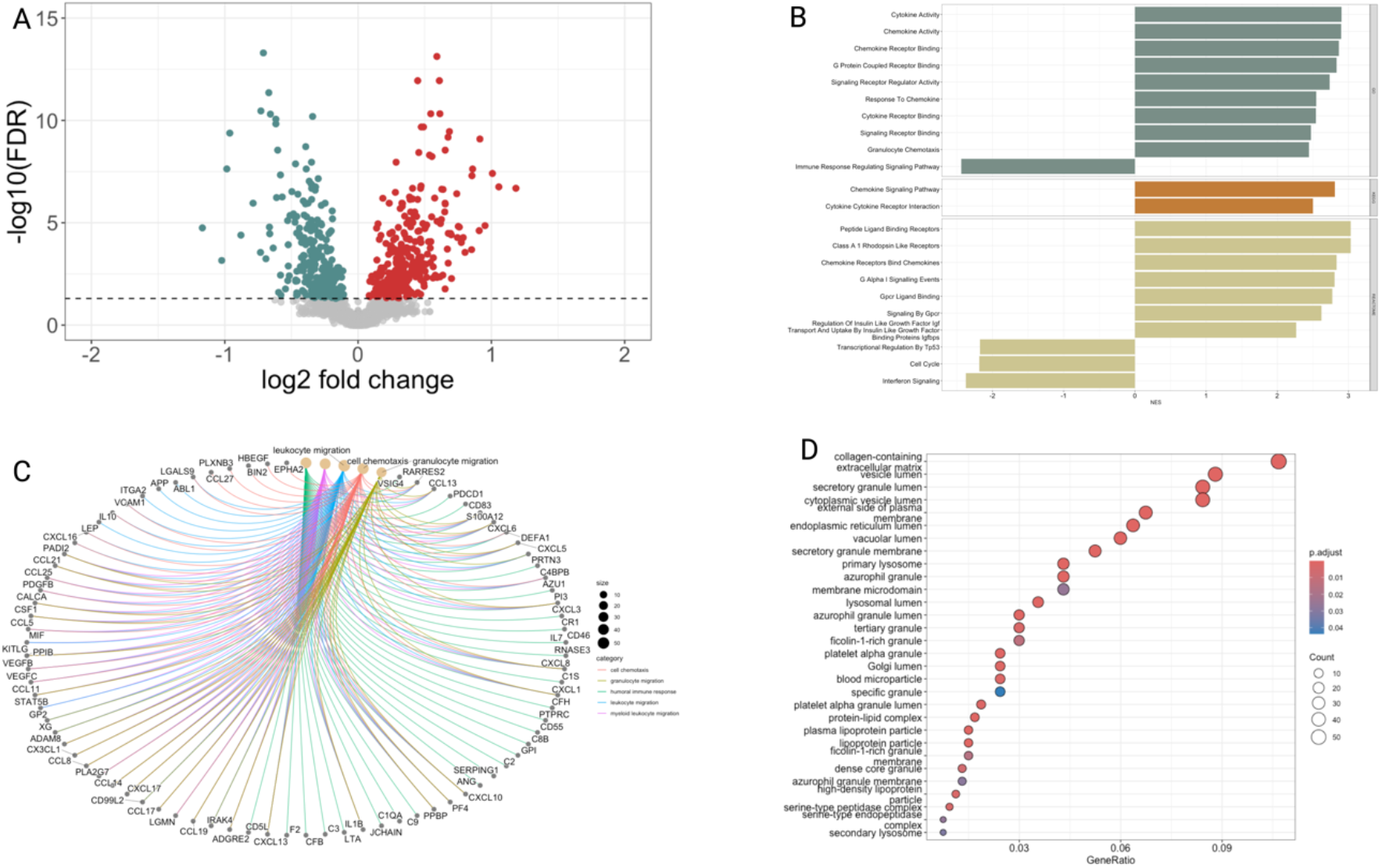
Volcano plot showing the differentially expressed proteins, with significantly overexpressed proteins annotated in red and downregulated proteins in teal using linear model implemented in Limma (A). Gene sets significantly associated with DEPs at 5% FDR (B). Biological process of significantly associated DEPs at 5% FDR(C). Cellular component of significantly associated DEPs at 5% FDR using a hypergeometric test (D

**Figure 3:**
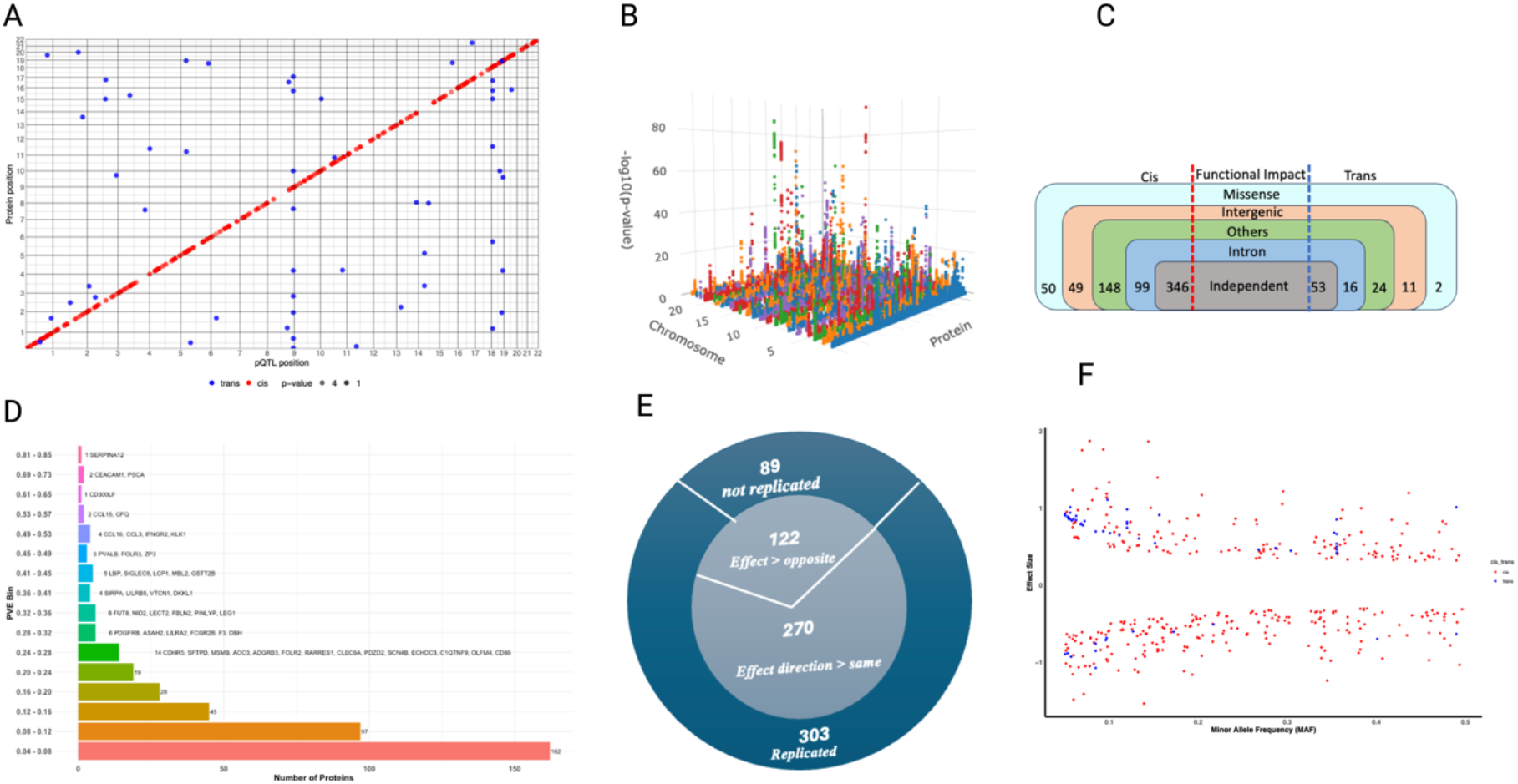
Scatterplot of pQTL variant location against the location of the gene encoding the target protein. Each dot represents an independent variant. Cis-pQTLs are colored in red, while trans-pQTLs are colored in blue (A). 3D Manhattan plot of identified pQTLs. (x axis = proteins, y axis = chromosome location, and z axis = −log10 p-values of each association) (B). Summary of identified pQTLs showing their functional consequences (C). Proportion of variance explained (PVE) by the conditionally independent pQTLs categorized into bins (D). Replication of conditionally independent pQTLs in the UKBB-PPP African-ancestry data highlighting replication via p values and direction of effect (E). Assessment of the relationship between the pQTLs effect sizes and minor allele frequency (MAF) (F).

We then undertook pQTL analysis with up to 15.8 million imputed variants for 2873 proteins, following quality control. Using approximate conditional and joint stepwise model selection, we identified 399 independent associations following multiple testing correction at P value thresholds of *P* < 1.46×10^−6^ and *P* < 2.2×10^−10^ for cis and trans pQTLs, respectively (supplementary 4). We identified 346 (86.7%) cis*-*pQTLs, and 53 (13.3%) trans*-*pQTLs. Seven proteins had both cis- and trans*-*pQTLs. We also identified 4 trans-pQTLs located within the following pleitropic loci: *PRSS27, CDH1, EPCAM, LPO, LEG1, TSPAN8, MUC2, SELE, IL7R, ALPI, KLK1, CDH17, CDH5, PTPRM*, and *SERPINI2*.

To determine the novelty of the pQTLs identified in the Ugandan population, we compared against pQTLs of 47 genome-wide pQTL studies (supplementary 5). We identified 6 independent cis-pQTLs and 31 independent trans-pQTLs that have not been previously reported in any population (supplementary 6). We compared our pQTL findings against the African-ancestry data of the UKBB-PPP and found that 16.7% (58 out of 346) of the discorvered cis-pQTLs and all trans-pQTLs have not been previously reported (supplementary 7). We tested the conditionally independent Uganda population pQTLs for replication in the UKBB-PPP. Of the 399 pQTLs, we were able to test 392 in the UKBB-PPP data. Of these, 303 replicated at P ≤ 1.2 × 10^−4^ (Bonferroni-corrected threshold) and 270 of these also had the same effect estimate direction (supplementary 8).

Next, we performed colocalization analysis to determine shared risk variants between the pQTLs and T2D using the largest multi-ancestry genome wide association study (GWAS) meta-analysis to date ^33^. We found evidence for a shared T2D risk variant (posterior probability 4 (PP4) = 95.5%) with pQTLs for the signal regulatory protein alpha (SIRPA) protein (Figure 4A,B). Integrating pQTLs with T2D GWAS revealed protein-disease colocalization that may suggest the involvement of SIRPA in the pathogenesis of T2D. SIRPA is a member of the SIRP family and is expressed on the surface of most T cells and some B cells. Genetic studies have implicated SIRP signaling in diabetes pathogenesis. For example, a SNP in human *SIRPγ*, encoding a SIRP family receptor that also binds CD47, was found to be associated with type 1 diabetes (T1D) ^34^. Wong *et al*., also identified SIRPA as a causal gene that mediates diabetes susceptibility in a nonobese diabetic mouse model ^35^.

**Figure 4:**
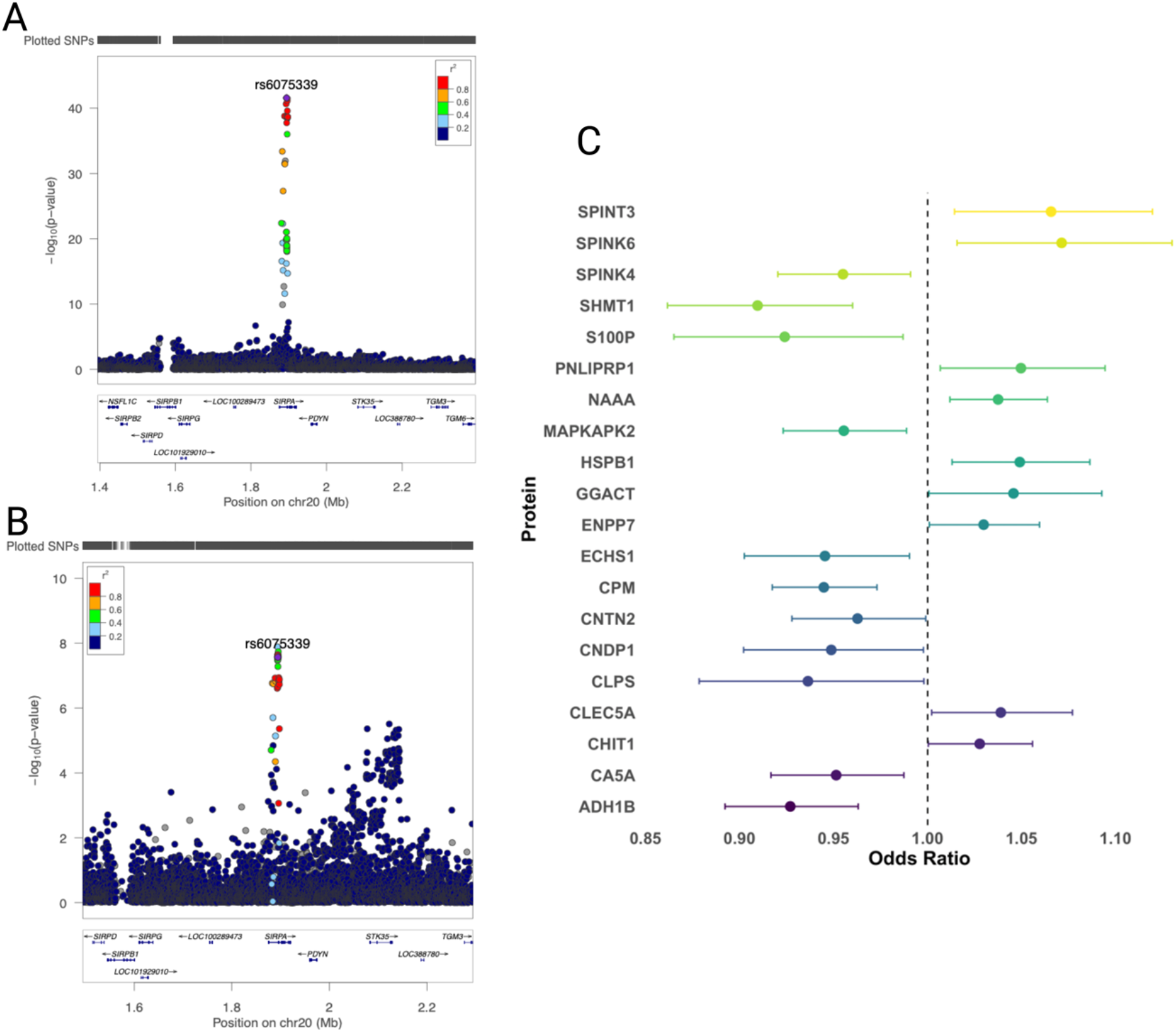
LocusZoom plots of the colocalizing SIRPA pQTL and T2D risk variant. The top panel shows the T2D GWAS p-values, while the bottom panel shows pQTL p-values for the same region (A,B). Mendelian randomization forest plot for proteins causally associated with T2D (C).

To examine the causal relationship between the identified cis-pQTLs and T2D, we undertook a Mendelian randomization analysis. We found 20 proteins to be causally associated with T2D. Higher levels of SPINT3, SPINK6, PNLIPRP1, NAAA, HSPB1, GGACT, ENPP7, CLEC5A, and CHIT1 were associated with increased risk of T2D, while SPINK4, SHMT1, S100P, MAPKAPK2, ECHS1, CPM, CNTN2, CNDP1, CLPS, CA5A, and ADH1B showed a protective effect on T2D risk (Figure 4C, supplementary 9). Several of these proteins such as PNLIPRP1, NAAA, HSPB1, ENPP7, CLEC5A, CHIT1, SHMT1, S100P, MAPKAPK2, ECHS1, CPM, CNDP1, CLPS, CA5A, and ADH1B have been shown to be associated with diabetes through pathways involving lipid metabolism, inflammation, protease activity, and hypermethlation etc ^36–39^. PheWAS analysis also showed that some of these proteins, such as SPINT3, SHMT1, ADH1B, and CNTN2, are associated with metabolic traits (Figure 5).

**Figure 5:**
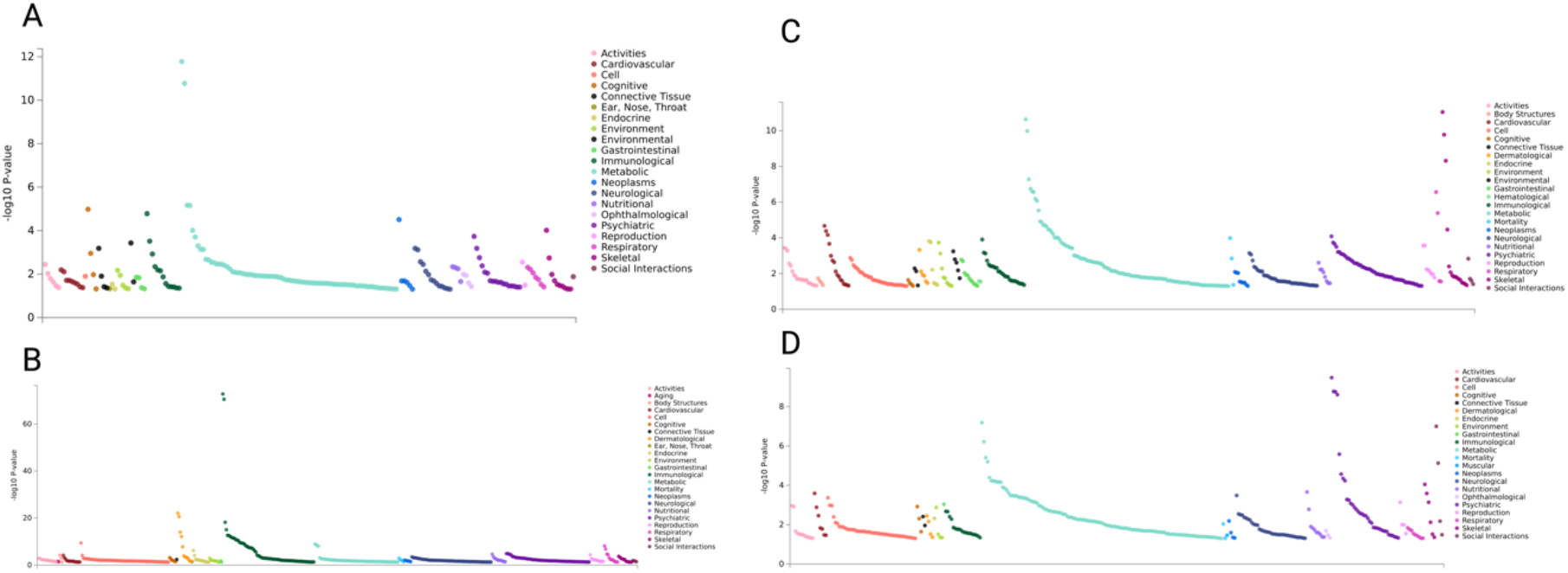
PheWAS plots for SPINT3 (A), CNTN2 (B), SHMT1 (3), and ADH1B (D).

Lastly, we assembled a list of 1804 postulated effector genes for T2D from 9 GWAS studies. If a gene coding for any of the proteins associated with identified pQTL in our study was found in the curated list, we defined such gene/protein as reported, if not, we classified them as previously unresolved. We identified 320 proteins previously unresolved as potentially linked to effector genes for T2D based on these GWAS signals (supplementary 10).

Our work takes a first step toward addressing the underrepresentation of continental African individuals in genetics and proteomics studies. As a result, we were able to delineate the molecular landscape of 2873 unique proteins in a context that might be pivotal to understanding drivers of T2D pathophysiology, identified 58 African ancestry-specific cis-pQTLs that have not been reported previously, and identified 20 proteins that are causally associated with T2D. The generalizability of these findings may be limited for the continent, since the population is drawn from a single demographic group within Africa. There is, hence, a need to include more ancestrally diverse populations in future studies.

Here, we have used the Olink targeted proteomic assay, which has some limitations, for example only a subset of the full proteome is studied, and the affinity of aptamers may be affected by missense variants. While HbA1c is a highly standardized and accurate test with lower intraindividual variability compared to fasting glucose, in African ancestry individuals using HbA1c as a blood sugar level indicator may not provide the full spectrum of the metabolic conditions associated with T2D due to the prevalence of hemoglobinopathies such as glucose-6-phosphate dehydrogenase (G6PD) deficiency. In individuals with G6PD deficiency, there is increased susceptibility to hemolysis, which may lead to reduced HbA1c level potentially leading to missed T2D diagnosis ^40,41^.

In conclusion, the novel associations and causally-associated proteins identified offer promising avenues for the development of targeted therapies and personalized treatment strategies for T2D, contributing to improved management and prevention of this global health challenge. Our findings demonstrate the utility and discovery opportunities afforded by the inclusion of African ancestry individuals in large-scale proteomic studies.

## Methods

### Study population

The participants were selected from the Uganda Genome Resource (UGR), which is a subset of the General Population Cohort (GPC). As previously described ^42,43^, the GPC is a population-based cohort made up of over 22,000 people from 25 nearby communities in the remote Southwest Ugandan sub-county of Kyamulibwa sub-county, which is a part of Kalungu district. We selected 528 samples from the UGR based on their age, sex and HbA1c. Following hemolysis of anticoagulated whole blood, the concentrations of total haemoglobin and HbA1c were measured using turbidimetric inhibition immunoassay-quant Haemoglobin Alc Gen. ^42^. In addition to the genotype QC described in ^42^, we used Hardy Weinberg p-value (pHWE) < 1×10^−6^.

### Association with clinical characteristics

We used linear regression to determine the association between the protein levels and systolic Systolic Blood Pressure (SBP), Diastolic Blood Pressure, alanine, albumin, alkaline, aspartate, bilirubin, cholesterol, gamma-glutamyl transferase, High-Density Lipoprotein, Low-Density Lipoprotein, triglycerides, and Hemoglobin A1c. All *p*-values were FDR corrected.

### Differential Expressed Proteins and Functional enrichment

We determined differential expressed proteins (DEP) between the cases and control using limma ^44^, and we used Benjamini–Hochberg False Discovery Rate (FDR) for multiple testing ^45^. DEP are defined as proteins with an FDR < 5% and a fold change greater than 0.5 (|log2FC| > 0.5). To better understand the functional impact of the proteins, we used enrichr tools from clusterProfiler ^46^ and then used Pearson’s correlation analysis to investigate the association between the expression levels of the DEPs. STRING database ^47^ was used to generate protein-protein interaction network for the upregulated and downregulated protein categories.

### Proteomics Quality Control

Olink’s proximity extension assay (PEA) technology ^48^ was used to measure the plasma level of 2978 proteins in 528 samples across eight Olink panels. The levels of protein expression were measured logarithmically as Normalized Protein eXpression (NPX) units. We adjusted all phenotypes using a linear regression for age, sex, plate number, sample collection season, followed by an inverse-normal transformation of the residuals. During the quality control (QC) process, we excluded one sample because the PCR plate well was empty, additional 2 samples were further excluded due to missingness greater than 40%. Two samples SC1 and SC2 were also excluded, these samples were initially included by O-link for internal control process. For assay QC, 40 assays were excluded as they did not have Normalized Protein eXpression (NPX) values. Additionally, we excluded 31 assays that had fraction of assay warning greater than 15%. No assay was excluded because of limit of detection (LOD). In all, 525 samples and 2873 assays remained after QC and were subsequently used for further analysis.

### Single-point association

Covariates such as sex, age, plate, and mean protein expression per sample were regressed using R’s LM function. The residuals were then translated into z-scores and used for association analysis. We used the single-point-analysis-pipeline version 0.0.2 (dev branch) [https://github.com/hmgu-itg/single-point-analysis-pipeline/tree/dev] to perform the association analysis. GCTA version 1.93.2 beta was used to conduct a mixed linear model association (MLMA) analysis, genetic relationship matrix (GRM) function within the GCTA software was used to estimate the genetic relationships among the individuals. We then used GCTA-COJO, which is designed for approximate conditional and joint stepwise model selection to identify independent associated variants at each locus.

### Significance threshold

Cis significant threshold was determined by first multiplying the Bayes Factors (BF) by 2875, and values over 1 were capped at 1. The BF was estimated using eigenMT ^49^. eigenMT calculates Meff as the number of ranked eigenvalues from the adjusted genotype correlation matrix needed to account for 99% of the detected genotype variability. Subsequently, the corrected p values were adjusted for multiple testing by applying the False Discovery Rate (FDR) method. Q-values were then calculated using the qvalue package, allowing for the identification of a subset of significant associations based on a q-value threshold of <0.05. Finally, the cis threshold for significance in pQTL analysis was determined by averaging the smallest non-significant p-value and the largest significant p-value. This method resulted in a cis *P*-value threshold of 1.462E^-06^. Trans threshold was calculated based on the effective number of variants (*N*_eff_) and number of protein traits (*M*_eff_). The *N*_eff_ was derived by performing LD-pruning with the following parameters indep 500 5 0.2 in Plink 1.9 ^50^. This resulted in an *N*_eff_ of 452593 unique variants. The *M*_eff_ was calculated using the *M*_eff_ function and Gao method in poolr R pckage ^51^. The trans *P*-value threshold is 2.227E^-10^. Variants within 1 megabase (Mb) upstream or downstream of the encoding genes are referred to as cis-pQTLs while trans-pQTLs are those found beyond 1mb relative to the encoding gene. Ensembl’s Variant Effect Predictor (VEP) was used to determine the functional impact of the variants.

### Comparison of pQTLs to prior published data

To determine the novelty of our pQTL, we used an in house-built database of previously identified signals of 46 genome-wide pQTL studies including the UKB-PPP ^15^. We evaluated novelty by identifying novel loci and novel variants. Novel loci were defined as those with no published variants within ±1Mb of our variants. For variants at known loci, we checked their rsIDs against those previously reported. Variants with no prior matches were further conditioned in the context of other known variants at that locus. These were classified as novel if the significance of their association p-value persisted even after adjusting for other known variants.

### Colocalisation analysis

We perform Bayesian-based colocalisation analysis using the Coloc.fast function (https://github.com/tobyjohnson/gtx) between our pQTL signals and multi-ancestry T2D GWAS summary statistics ^33^ from DIAGRAM database. To assume shared genetics, we used default priors and posterior probability of (PP.H4) ≥ 0.8 ^52^.

### Mendelian Randomization

We used the TwoSampleMR package ^53^ to undertake an MR analysis between the proteins and African ancestry T2D to further determine causality and the direction of effect. Wald ratio test was conducted for proteins with a single cis-pQTL variant and IVW was used as the main analysis in instances with more than 1 variants.

### Identification of Effector Genes

To find putative effector genes for T2D, we compiled effector genes associated with T2D GWAS. This dataset was curated from nine papers published in the Type 2 Diabetes Knowledge Portal (T2DKP), resulting in a collection of 1,804 distinct effector genes. For classification purposes, proteins that were documented in our curated list were labelled “reported.” Those not found on the list were classified as “unresolved.”

## Data Availability

All data produced in the present study are available upon reasonable request to the authors

## Ethics

The study was approved by the Uganda Virus Research Institute Research and Ethics Committee (UVRI REC #GC/127/907) and the Uganda National Council for Science and Technology (UNCST HS2527ES).

## Code Availability

Analyses were performed using publicly available software

## Acknowledgement

We acknowledge the support of the Core Facility-Metabolomics and Proteomics at Helmholtz Munich. We thank the core facility for help with sample preparation and protein measurement. We thank all participants who contributed to the Uganda Genome Resource. UGR/GPC was supported by the UK Medical Research Council (MRC) and the UK Department for International Development (DFID) under the MRC/DFID Concordat agreement, through core funding to the MRC/UVRI and LSHTM Uganda Research Unit. The 2023 Award Fellowship support of the Alexander Von Humboldt Stiftung Foundation to OS is acknowledged. SF was supported by the Wellcome Trust grant number 220740/Z/20/Z.

